# Co-establishing an infrastructure for routine data collection to address disparities in infant mortality: Planning and implementation

**DOI:** 10.1101/2021.05.15.21257216

**Authors:** Naleef Fareed, Christine Swoboda, Tyler Griesenbrock, John Lawrence, Timothy R. Huerta

## Abstract

**Background:** Efforts to address infant mortality disparities in Ohio have historically been adversely affected by the lack of consistent data collection and infrastructure across the community-based organizations performing front-line work with expectant mothers, and there is no established template for implementing such systems in the context of diverse technological capacities and varying data collection magnitude among participating organizations.

**Methods:** Taking into account both the needs and limitations of participating community-based organizations, we created a data collection infrastructure that was refined by feedback from sponsors and the organizations to serve as both a solution to their existing needs and a template for future efforts in other settings.

**Results:** By standardizing the collected data elements across participating organizations, integration on a scale large enough to detect changes in a rare outcome such as infant mortality was made possible. Datasets generated through the use of the established infrastructure were robust enough to be matched with other records, such as Medicaid and birth records, to allow more extensive analysis.

**Conclusion:** While a consistent data collection infrastructure across multiple organizations does require buy-in at the organizational level, especially among participants with little to no existing data collection experience, an approach that relies on an understanding of existing barriers, iterative development, and feedback from sponsors and participants can lead to better coordination and sharing of information when addressing health concerns that individual organizations may struggle to quantify alone.

## Background

### Introduction

The use of community-based organizations to address social determinants of health has a long history [1]. Infant mortality represents one area where such an approach has been of common use [1-4]. Ohio is ranked as the ninth-worst state in the United States for infant mortality in 2019, with an estimated rate of 7.3 infant deaths per 1,000 births [5]. Further, there has been significant published research and numerous reports that have identified the notable disparity in infant mortality between white and black infants; the infant mortality rate is almost three times higher among black infants (13.9 per 1,000 births for black infants in 2018 vs. 5.4 for white infants) [6-11].

In 2012, the state legislature renewed efforts to address this disparity by convening local partners in the nine counties in Ohio that account for the majority of black infant mortality. In addition, state agencies collaborated to use population data to target areas for outreach and services as well as coordinate efforts among the multiple programs in each community. Infant vitality efforts in these nine counties, also known as the Ohio Equity Institute (OEI) communities [12], include group-facilitated prenatal care, home visiting, progesterone administration, breastfeeding support, fatherhood programs, safe sleep, and health education and community engagement activities focused on changes such as tobacco cessation and birth spacing. Coordinating these efforts helped bring focus to the number of independent initiatives to improve infant and maternal health; however, the effort also highlighted an absence of consistent data collection and infrastructure across programs to facilitate evaluation.

In 2017, the Ohio Department of Medicaid (ODM) began to focus on the implementation of a consistent data collection and infrastructure across programs to facilitate evaluation despite access to Medicaid and population data. The vision refocused on using data to increase surveillance to target areas for outreach and services as well as the coordination of efforts among the multiple programs in each community. To address the lack of consistency across data collection, ODM developed the OEI Community-Based Organization (CBO) Evaluation project. The goal of the OEI CBO Evaluation project is to create a statewide system for routine data collection and conduct analysis on the data collected to determine the extent to which the selected interventions serve high-risk pregnant women and assess the effect of these interventions on health care utilization and birth outcomes. The research team is made up of researchers at The Ohio State University and the Ohio Department of Medicaid, both in Columbus, Ohio. Program managers attended meetings in person, online, or via phone with the research team in Columbus. The Ohio State team, composed of health and bioinformatics subject matter experts, was tasked with developing a multimodal data collection system.

At the start of the evaluation, the research team performed a needs assessment, which noted there was significant heterogeneity among the interventions, at times individuals participated in more than one program, and there was no standard for data collection. Without a multimodal data collection model necessary to support public health and surveillance of infant mortality, there was a great deal of disparity in data collection approaches and infrastructure. Further, there were no standards for reporting such community-based data. This paper describes the multimodal data collection model necessary to support public health and surveillance of infant mortality and serves as an exemplar for how such efforts might be accomplished in other domains.

Taken together, concerns about the disparities in the existing approaches to data collection and the associated impact precipitated into a call to action by state agencies to develop a robust data collection infrastructure for OEI. The primary objectives of developing our data model for OEI were twofold: 1) to generate reports that quantify the performance of the CBOs funded by OEI based on key metrics and 2) to dynamically visualize these key metrics using a Tableau dashboard.

#### Prior work

As part of this project, we sought to design a data model that included characteristics we previously determined to support public health and surveillance of infant mortality in Ohio. Active data collection processes have been shown to improve prevention programs and increase evaluative capacity to address multiple public health problems [13-17]. A robust data collection system for OEI was determined to have the following characteristics: 1) multimodality of data collection, 2) a standardized data model, and 3) systematic educational outreach efforts to train those reporting data.

A multimodal system is one that allows for data collection and delivery across a number of communication channels: phone, email, fax, mail, and direct data entry. Multimodal systems allow for data collection across CBOs that vary in their technological capabilities and resources. CBO staff have varying levels of comfort with using tools to report data; some are more comfortable using information technology, while others gravitate to using paper forms. The final data is electronic, and paper forms have additional data entry and submission steps; however, many of the community organizations do not have the technology to collect data online, and it may be easier to collect data on paper during in-person visits with participants. Prior research indicates improved data quality when multiple data collection modes are available [17-20].

A standardized data model allows for the collection of a common set of core metrics across CBOs with heterogenous activities and data collection practices, improving cross-program comparability [21]. The wide variation in the data collection among CBOs before the OEI CBO Evaluation project may have created challenges that were difficult for CBOs such as not directly targeting key interventions. Some CBOs may not have collected enough data to determine whether their participants were high risk, identify service provision needs, or evaluate program efficacy. It has been noted that instituting such a model, also known as a common data model (CDM), is challenging, especially with a diverse set of health care organizations that have varying levels of requisite resources to populate the CDM [22]. Notwithstanding, a CDM would enable new research opportunities that help with the improvement of interventions and assessment with outcomes that are based on data that were previously not available or limited to single site studies [23].

Systematic educational outreach efforts are necessary to train health care providers on how to report data to a CDM, maintain the fidelity of data collection over time, and foster relationships between community health organizations and research teams [17, 24-26]. One study demonstrated that use of training elements and data trainers allowed for public health workers to have better awareness of data reporting and higher data reporting rates than those who did not have such resources [25]. Another study listed the early misperceptions associated with routine data reporting – which included providers not realizing the value of data reporting, the inability to fit data reporting with existing workflows, and the lack of best practices on reporting data – and noted how these views diminished over time with the support of training and guidelines [24]. Effective public health surveillance requires training covering key concepts, terminology, policies, information technology, and practices related to data reporting [17].

#### Goal of this effort

This paper describes the design, development, deployment, and assessment of a data collection system to improve public health efforts by collecting data from multiple community programs. The goals of our effort were to minimize data collection burden while maximizing the ability to engage in evaluative assessment of the effect of participation in OEI CBOs. Our paper serves as a template for the development of similar systems, in other contexts and other locations, in which existing data infrastructure and inter-organizational coordination is insufficient to meet the needs of a public health effort. We describe the design of the OEI data collection system and data model that supports these goals. The system developed is still being used to collect data from CBOs across the state for the Ohio Department of Medicaid.

## Methods

### Study population

The OEI CBO Evaluation project focused on programs in nine counties in Ohio with large disparities in infant mortality and significant urban populations: Butler, Cuyahoga, Franklin, Hamilton, Lucas, Mahoning, Montgomery, Stark, and Summit. These include the urban areas in the cities of Hamilton, Cleveland, Columbus, Cincinnati, Toledo, Youngstown, Dayton, Canton, and Akron, respectively. Existing and newly developed community programs received funding to provide consistent data to ODM and could use this funding for hiring, training, and general program needs.

### Prototyping the OEI data collection infrastructure: Needs assessment and design

The research team at The Ohio State University for OEI consisted of faculty, skilled professionals, and technical experts tasked to develop, deploy, and evaluate the data collection system. First, in early 2018, our team conducted a needs assessment by reviewing previous program data collection materials, program needs identified during interviews and focus groups with CBOs, and data collection material from similar programs identified during literature review to conceptualize a multimodal data collection system. Based on our engagements with the CBOs, the research team sought to sketch out for the communities what a robust infrastructure might entail. The needs assessment occurred over a two-month period.

Our needs assessment revealed that the CBOs that collect the most data have data collection activities at intake; each time a participant is seen; and, for some programs, after birth and when exiting the program. Data collection is done either during home visits with a participant, at group classes, or in other one-on-one settings such as public offices. Some programs collect most of the intake data at the first visit, while some collect it over a few initial visits. Our OEI data collection system largely mimics this, having forms for intake, birth, exit, and encounter with online and paper data collection options.

One of the major gaps noted by our needs assessment process was that without a system of this nature, there was excess variation in data collection approaches between CBOs. For example, initial engagement identified some community programs with advanced electronic data collection systems in place, while other CBOs worked entirely with paper records. We found that some CBOs collected only aggregated attendance data, while others maintained databases of extensive medical, behavioral, environmental, and demographic data. In addition, sufficient identifier information was often not collected, precluding post-processing and subsequent matching of data with community infrastructure.

The needs assessment revealed potential benefits of implementing a more robust system: the sponsors would receive more consistent data collection and program evaluation, while CBOs collecting minimal data would receive data collection materials and support to help with evaluation. Most CBOs had performed little evaluation work; employees lacked the time and resources, and the numbers of participants did not allow for robust analysis of outcomes. The absence of this system could be linked to implications for CBOs such as potential suboptimal allocation of resources, not targeting the individuals who may benefit most, and not providing as many referrals or interventions as a participant may need due to being unaware of risks.

The research team developed an initial variable list with the sponsors that was revised based on the needs assessment. Our initial variable list included variables measuring demographic data, environmental and behavioral risk data, and data about care received by the mother and infant. The demographic data collected included names, birth dates, and Medicaid or Social Security identification numbers to ensure that Vital Statistics and Medicaid data are matched properly, as well as address information to map where participants reside. The needs assessment revealed a desire from the CBOs to not have an overly burdensome number of data points. Most already collected much of the data and would not be able to significantly add more data points. Many clinical variables and birth-related variables that can be found in other materials were removed from the variable list to reduce data collection burden from participants. Many of the variables not collected are linked into the final dataset from Vital Statistics birth records, including birth weight, gestation at birth, and other health variables. Data collection materials were developed once the variable list was finalized. These variables would subsequently be collected and curated on a database server that concomitantly allowed for the querying of data through of a common data model. All data collection activities were approved by an Institutional Review Board at the Ohio Department of Medicaid.

Figure 1 illustrates our vision for the OEI data collection infrastructure. The figure also shows how the various data collection, curation, and reporting activities are integrated within the infrastructure system. The first step of the system involves a CBO provider collecting information about a participant using one of the previously described collection forms (i.e., intake or birth). The subsequent step is for the provider to use a data entry mechanism to report the data to the research team. Transfer of data is possible through an online data entry system, scanning and faxing forms, mailing forms, secure email of spreadsheet data or forms, or uploading to a secure online portal; in our case, we have used the Qualtrics platform, which accepts data both input into a survey or securely uploaded as a file. The third step involves the curation of a database by the research team on a server that, in step four, can be used by the team to access data via a portal. Step five illustrates how the research team can use the curated database to develop a CDM that can be used by researchers and government agencies to develop queries for reports and dashboards. The CBOs can also use this data to interact with the researchers and government agencies for decision-making purposes. Challenges with character recognition applications led to the use of manual data entry.

**Figure 1.** Vision for the Ohio Equity Initiative data collection infrastructure.

### Development of the OEI data infrastructure and common data model

Between February 2018 and July 2018, our team engaged in the development and deployment of the OEI data collection infrastructure. Critical early milestones during this period were developing an agreed upon timeline for data reporting, piloting data collection forms with select CBOs, and refinement of these forms. Subsequent milestones focused on piloting the data collection system between July 2018 and August 2018 across all CBOs, testing system integration, validating data points, and making system refinements based on the feedback obtained from the previously listed activities. Data collection began in September 2018 for 30 programs, and additional programs were enrolled over time after training and data use agreements were finalized. The data use agreements allowed the organizations to share individualized data with the research team that is aggregated and evaluated before being presented to the sponsors or other programs.

During the development period of the data infrastructure, our team concurrently focused on the design considerations and development of the OEI common data model. The data model architecture for our project contains data about the OEI program, its participants, and non-OEI participants. The model consists of elements from our data collection infrastructure (i.e., OEI system) linked to data sets from state databases (i.e., Ohio Vital Statistics and Medicaid Claims). The online data collection system consists of a database using Qualtrics, which offers the ability to collect data about multiple participants, and from multiple CBOs. CBO members create login information and are directed to a separate landing page for each CBO. On this landing page, participant demographic information can be added and surveys can be filled out and updated about each participant. These Qualtrics surveys follow the same format as the other data collection methods; CBOs create a record of a participant, then have the ability to complete intake, birth, exit, and encounter forms for that participant. These reports are downloaded by the researcher directly and processed and appended. CBOs can also securely send an Excel-readable spreadsheet that is imported and processed, or scanned paper forms that require an additional data entry step into a spreadsheet that is later imported. Another layer to the collection of data through the OEI system is the submission by some CBOs of data formatted for other data collection systems, in our case Care Coordination Systems (CCS) and the Ohio Comprehensive Home Visiting Integrated Data System (OCHIDS). CCS and OCHIDS contain similar data to the OEI system and required some standardizing before appending to the master dataset.

### Training to use the OEI data collection infrastructure

CBOs were trained on how to use the data collection materials in July and August 2018. These training sessions consisted of webinars with demonstrations of how to use the Qualtrics data portal, the validated Excel spreadsheet, and the paper forms. After these demonstrations, CBOs were given a choice of how to submit data between those options. After this choice, the CBO was provided with paper forms, the spreadsheet file, or login information to start data collection. After the first month of data collection, surveys and phone calls were conducted to discuss this process. CBOs were given the opportunity to change data submission preferences, provide input about portal changes desired, and receive additional training.

## Results

### Data elements

Data collection occurs at four points in time: enrollment, encounter, birth, and exit. Supplementary Table 1 in Additional File 1 lists the variables from the enrollment form. The intent of the enrollment form is to collect baseline demographic, risk factor, and history of prenatal care information. Supplementary Table 2 in Additional File 1 lists information collected during an encounter with a provider. The intent of the encounter form is to primarily obtain attendance and service utilization information.

Supplementary Table 3 in Additional File 1 lists information gathered about the participant and their newborn infant. The intent of the birth form is to collect risk factor information related to the baby and care utilization by the mother. Birth record data is linked to program data, enabling the researchers to know about infant and maternal health outcomes without overburdening the participant with questions that are personal or may be self-reported inaccurately. Supplementary Table 4 in Additional File 1 lists information collected from the exit form. The intent of the exit form is to collect information about care utilization by the mother and infant. Use of resources from various public programs and risks factor information are also collected. A detailed code book for all the variables collected in the OEI data collection infrastructure is provided in the appendix.

### Data collection infrastructure

To develop the OEI system and integrate our data model into the system, we accounted for five critical constraints:

1. Data is currently gathered by CBOs for other purposes. To the extent that the CBO leverages its existing infrastructure to gather this data, we believe that data quality will be higher.
2. Some CBOs may not currently collect data. In these cases, we provided tools that facilitate data entry in a manner that reduces the likelihood of data integrity issues.
3. Some CBOs may not have the capacity to collect data in the local setting. We make no presumption of computational ability in these settings and facilitated data collection in a manner that allows the CBO to meet the reporting criteria with a minimum transaction cost.
4. Data collection is not the focus of these CBOs. Given the community nature of the interventions, the system was developed in a manner that supports ease of use.
5. Data may be collected within a CBO or at a participant’s home. The nature and timing of how CBOs are administered required a data collection approach that is robust across multiple settings.

Given those constraints, we developed our multipronged, multimodal data collection and

integration approach for the OEI system.

#### Data collection and integration use cases

The following section outlines data integration use cases and explicates how data was integrated in a single database.

##### Use Case 1: The CBO keeps digital records, collects all required data elements, and those elements conform to the data model specifications (Figures 2 and 3)

| **Figure 2** |  |
| --- | --- |
| Data Collection Workflow: | • The CBO collects data on its participant that includes all the required data elements.<br>• It maintains those records in digital form.<br>• It is able to extract data on those records digitally in .CSV, .XLS, .XLSX, or any other Excel-readable file. |
| Organizational Workflow: | • The CBO extracts a dataset and submits that data through either email or a web portal in Qualtrics. |

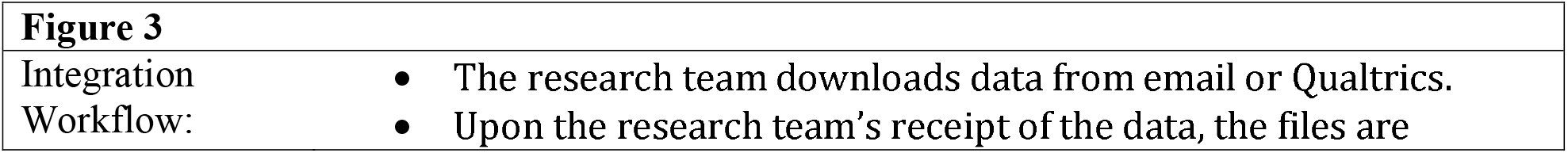
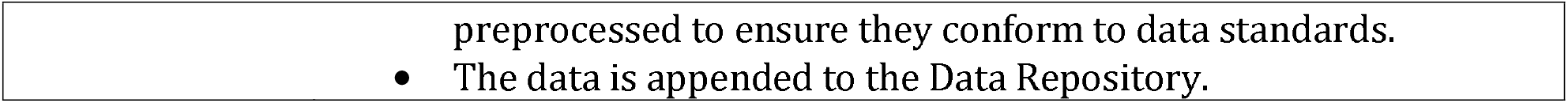

**Figure 2.**
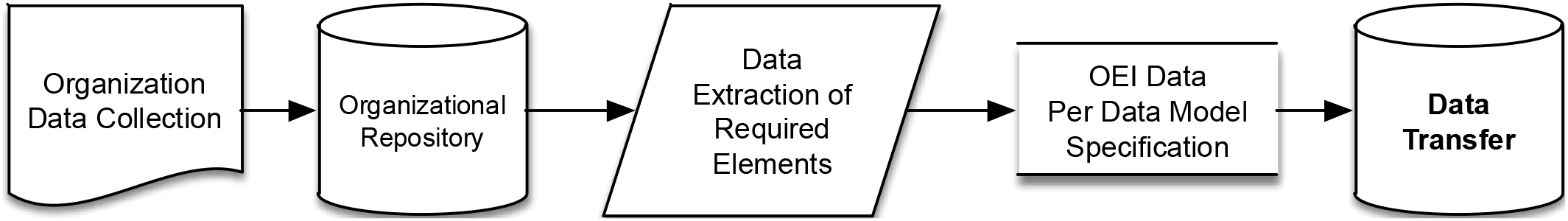

**Figure 3.**
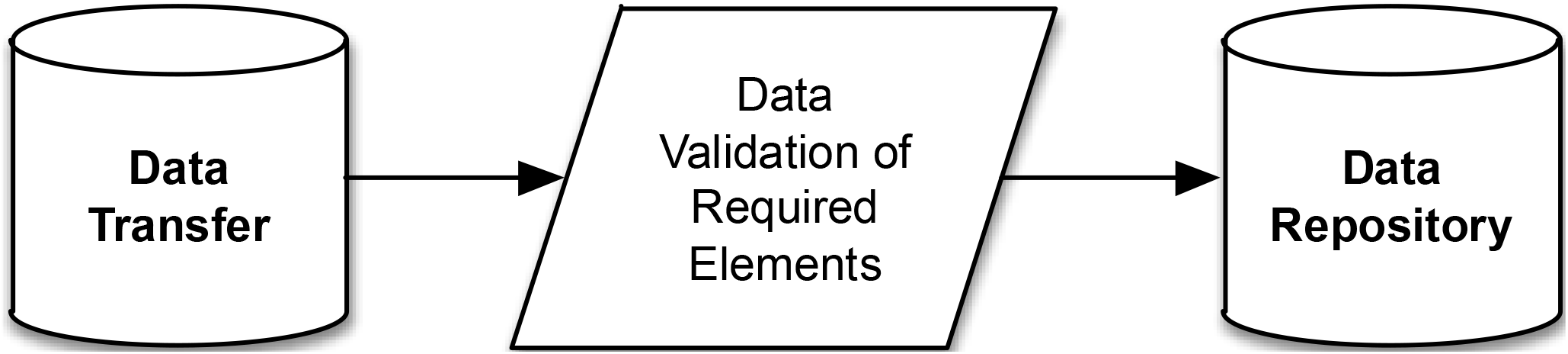

##### Use Case 2: The CBO keeps digital records but does not collect data that conforms to the data model (Figures 4 and 5)

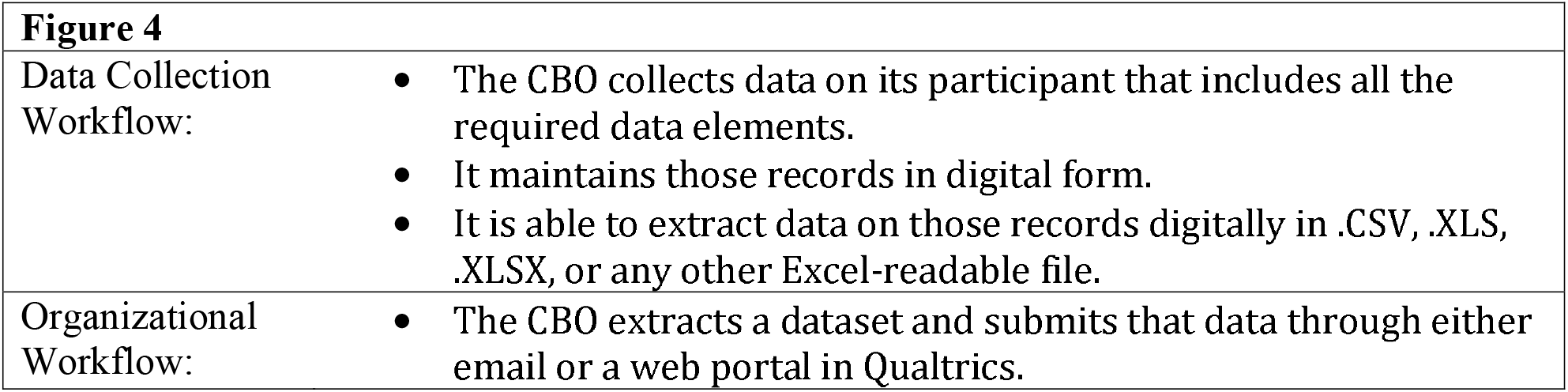

**Figure 5**
|  |  |
| --- | --- |
| Integration Workflow: | The research team downloads data from email or Qualtrics.<br>Upon the research team's receipt of the data, the files are preprocessed to ensure that they conform to data standards.<br>The research team uses a transformation script that maps the provided data transfer to the Data Model Specification.<br>The data is appended to the Data Repository. |

**Figure 4.**
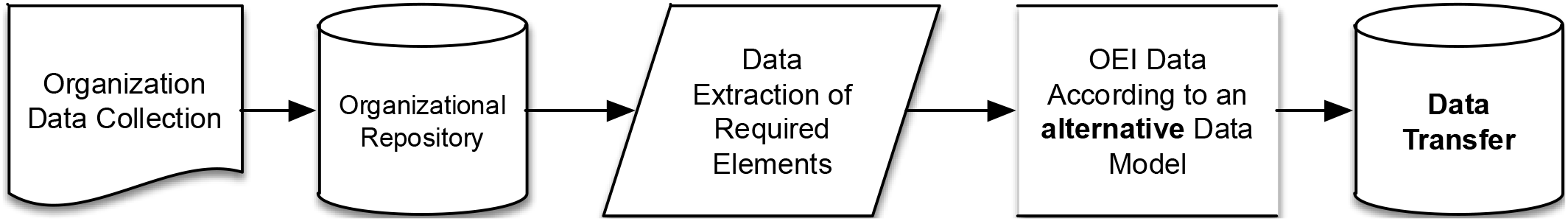

**Figure 5.**
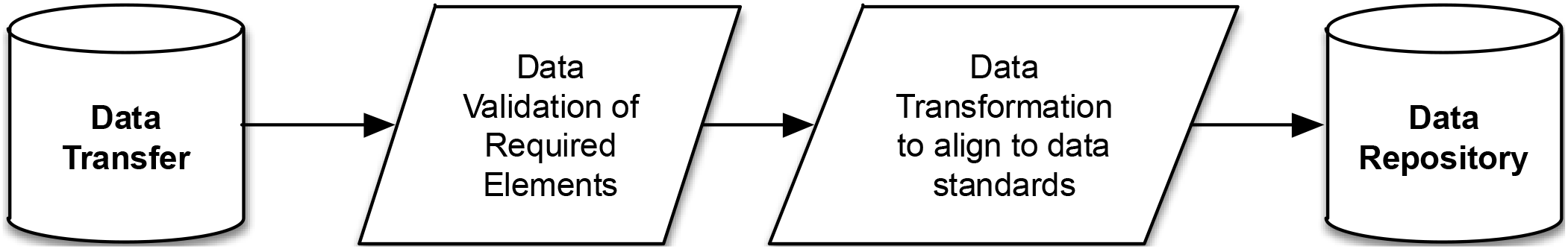

##### Use Case 3: The CBO keeps digital records, but it does not collect all required data elements and those elements do not conform to the data model specifications

In these cases, the CBO modifies the workflow in one of two ways:

1. Modify current systems to gather the additional required information and then conform to either Use Case 1 or 2.
2. Adopt the use of digital or paper forms and associated workflows.

For cases in which a CBO does not keep digital records, it chooses one of three options:

1. Use an online data submission system that mimics the paper forms.
2. Use a paper form and send it to our research team for processing.
3. Use a validated Excel spreadsheet created by our research team that mimics one of the forms from one of the other collection modalities.

##### Use Case 4A: The CBO does not currently keep digital records and chooses to use an online data submission system (Figure 6)

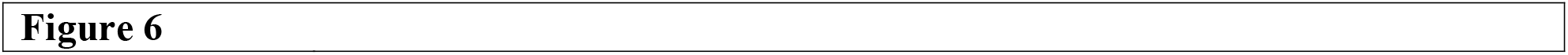
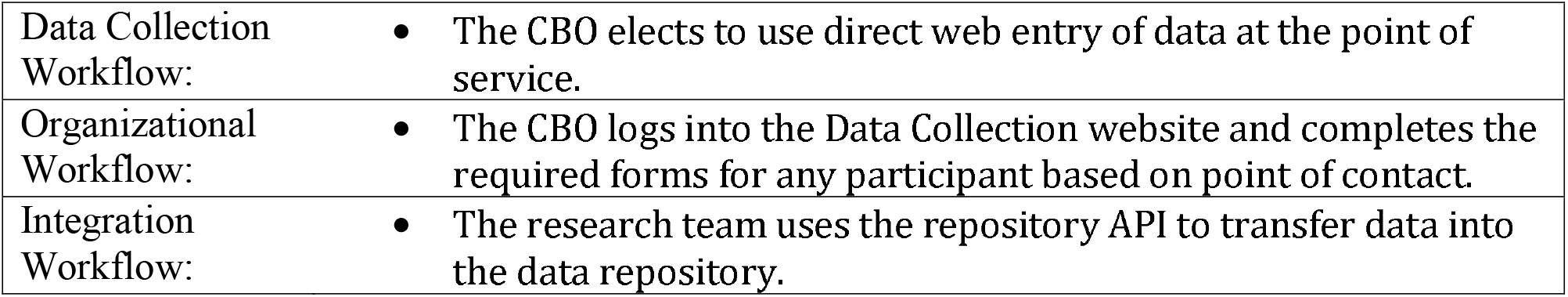

**Figure 6**
|  |  |
| --- | --- |
| Data Collection Workflow: | • The CBO elects to use direct web entry of data at the point of service. |
| Organizational Workflow: | • The CBO logs into the Data Collection website and completes the required forms for any participant based on point of contact. |
| Integration Workflow: | • The research team uses the repository API to transfer data into the data repository. |

**Figure 6.**
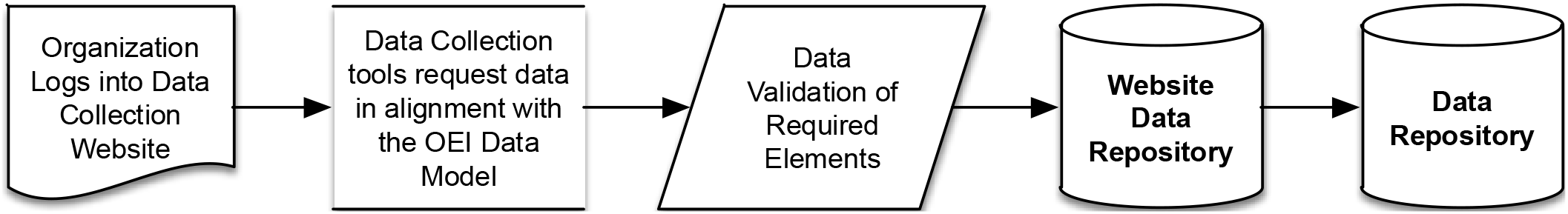

We provided an online system that allowed the CBO to enter data. The system required a computer with an internet connection. The website required site-specific authentication, and we provided credentials to the CBOs that chose this model. In cases when this approach is used, the workflow looks as follows:

##### Use Case 4B: The CBO does not currently keep digital records and chooses to use a paper-based data collection system (Figure 7)

**Figure 7.**
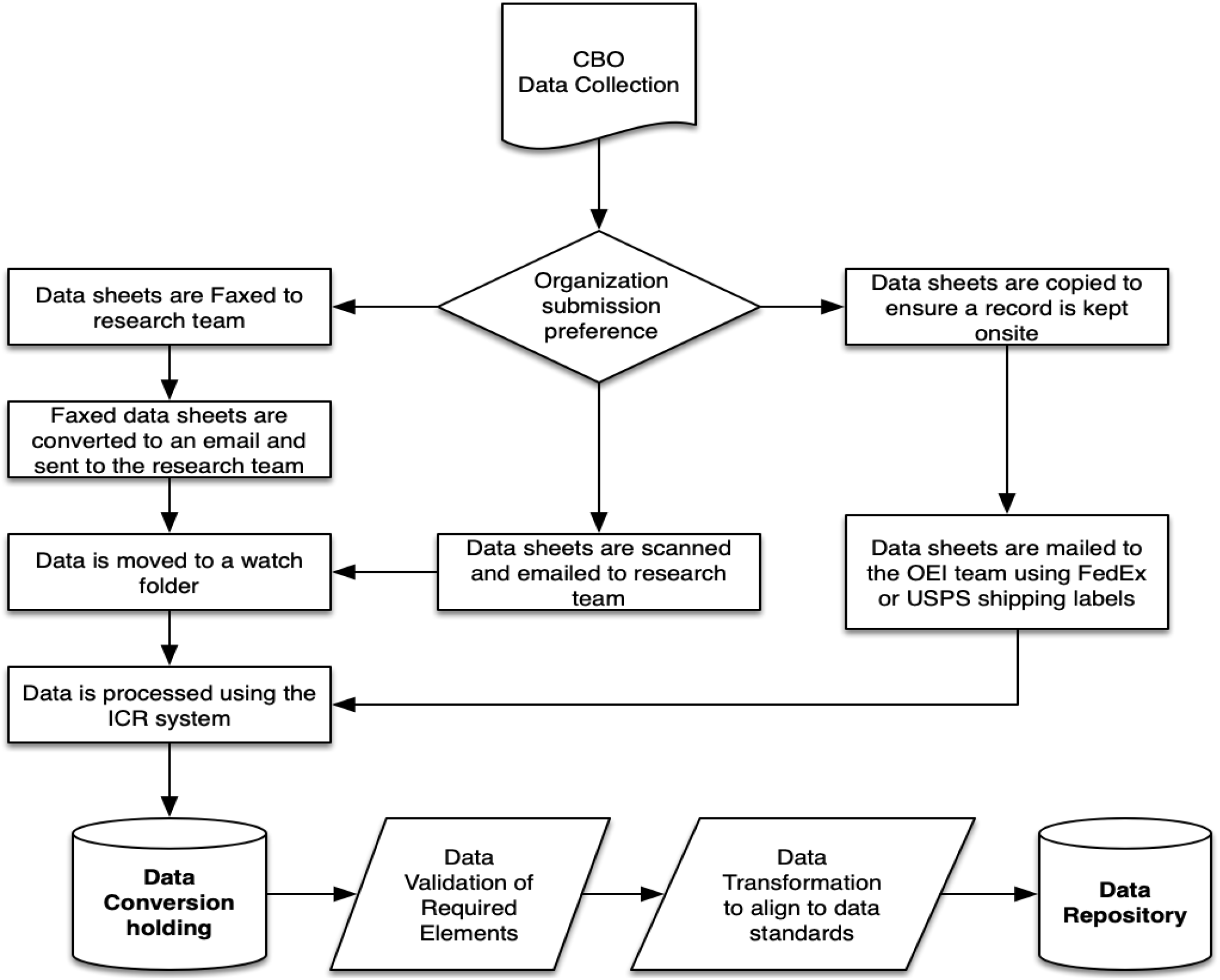

In the case when a CBO chooses to collect data on paper, we provided the CBO paper forms, and their digital equivalent, which they used to meet their reporting obligations. The forms are 8.5 by 11 inches, double sided, and singly designed to be used in all cases. CBOs using paper forms could also choose to adopt the model of Use Case 4A and enter data after the fact. In these cases, our team followed the workflow illustrated below:

|  |  |
| --- | --- |
| **Figure 7** |  |
| Data Collection Workflow: | • The CBO collects data on its participant that includes all the required data elements.<br>• It maintains those records in digital form (applicable to case 4A).<br>• It is able to fax, scan, or mail records to Ohio State (applicable to case 4B).<br>• It is able to extract data on those records digitally in .CSV, .XLS, .XLSX, or any other Excel-readable file. |
| Organizational Workflow: | • The CBO submits that data through fax, email, or mail. |
| Integration Workflow: | • Upon the research team's receipt of the data, the files are preprocessed to ensure they conform to data standards.<br>• The research team uses a transformation script that maps the provided data transfer to the Data Model Specification.<br>• The data is appended to the Data Repository. |

#### Database integration and analysis

Starting in October 2018, CBOs were asked to submit their previous month’s data by the 10th day of each month. Data from September 2018 was reported for 41 CBOs, and the number of CBOs reporting data increased to 62 by June 2019. From August 1, 2018, to June 1, 2019, the total number of participants reached across all programs was 10,693, representing 10,074 different people, indicating 619 clients participated in more than one program. Our research team received data from the CBOs and engaged in extensive data processing steps. These involved assessing data validity, accuracy, completeness, consistency, and uniformity. We also inspected the data for duplication of participant information, where we retained the most complete or most recent version of the forms. Once these steps were complete and the data sets were standardized, they were appended to one another. This appended data set was then matched to birth records and Medicaid records using key identifiers that included the participant’s Medicaid ID, name, and address; non-matching records were retained for our control group. Of the 8,984 total participants in the dataset used to match to birth records in May 2019, there were 2,829 participants (31.5%) matched to birth records and 5,798 participants (64.5%) matched to Medicaid records. Many of the participants had not yet given birth at the time of this matching. All the data sets in the database contained a one- to three-month delay after an event, except for the Medicaid claims data, which has a six-month delay.

Our research team continuously updated a data repository with the above described data that were appended with information from the OEI system and the external state data sets. Periodic extracts from the data repository were used to generate reports for key stakeholders (e.g., annual reports to ODM) and used in the Tableau Dashboard to visualize key metrics. The dashboards, built by the research team with regular feedback from stakeholders, display the data in the form of bar charts, pie charts, and tables of relevant information. Each page of the dashboard is able to focus on a different portion of the data, such as the enrollment numbers for the CBOs, gestation data, or risk factors. Making selections within the dashboard modifies the view and presents additional data. By offering the information in a more visual manner, rather than in tabular format, it can be put to use by policymakers and others who may not be as familiar with statistical analysis while allowing them to draw their own conclusions. An example of one of the OEI dashboards is pictured in Figure 8. (Synthetic data is used in the illustration.)

**Figure 8.**
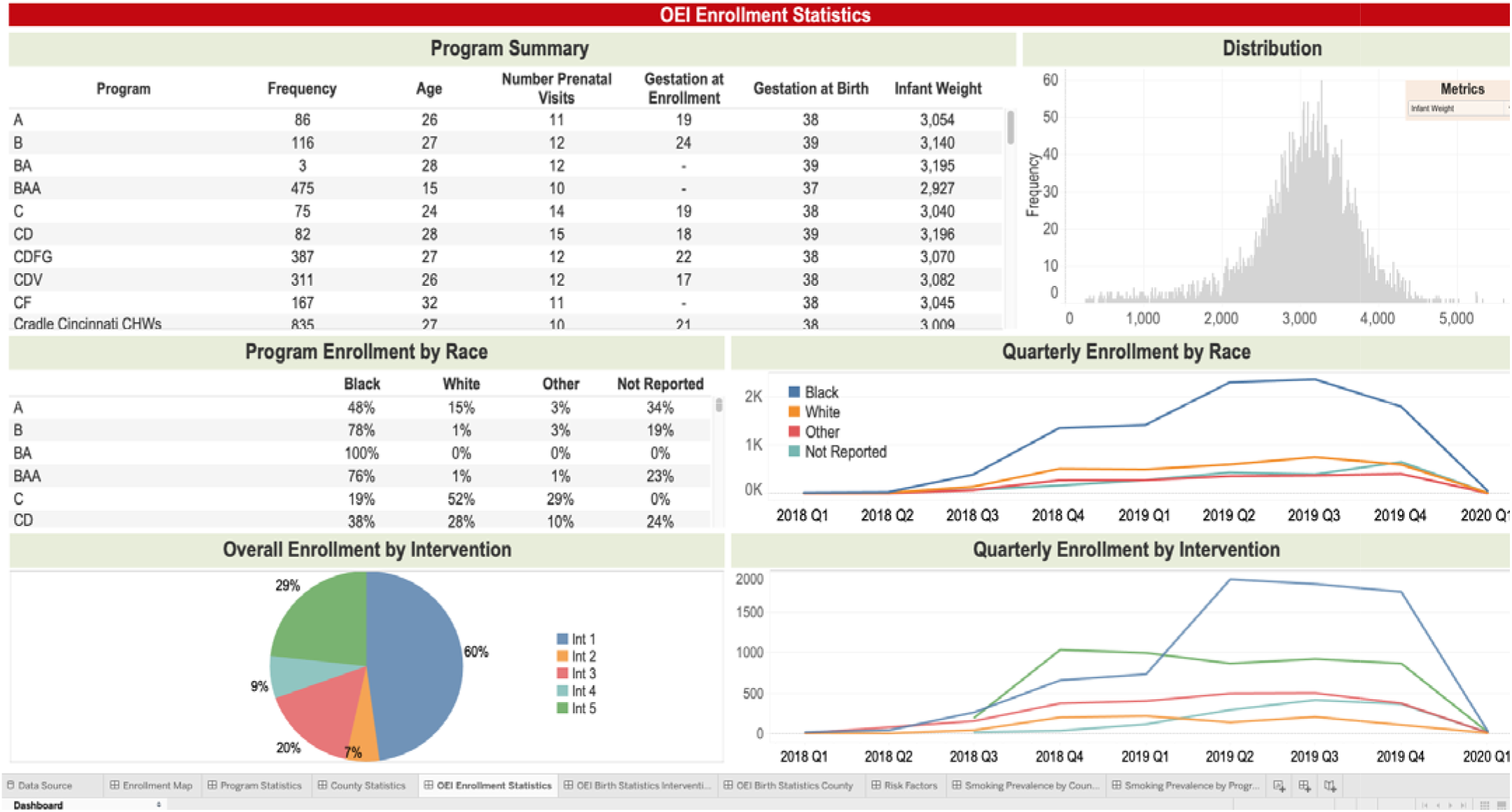
**This image from the OEI dashboard shows enrollment statistics for participating CBOs with details based on race, enrollment date, and other variables collected by the CBOs.**

#### Data quality

During the data processing step, our research team performed assessment of missingness of data for each variable measured. Overall and individual CBOs’ reporting of values for variables were inspected. CBOs that had poor data quality were contacted by our research team during one of multiple Plan-Do-Study-Act (PDSA) cycles to identify systematic barriers to data reporting. During our PDSA calls with the CBOs, our research team used this information to recommend solutions to help improve data reporting. We experienced notable improvements after each PDSA cycle, and this helped improve data reporting methods over the aggregate reporting period. For example, two CBOs that reported 5 out of 8 (63%) of the contact information variables in January 2019 were reporting all eight of the variables by May 2019. The total dataset had 95.0% of participants’ first names and 94.8% of their last names in January of 2019, and 99.3% of first names and 99.2% of last names by May 2019. In January, there were 31 programs sending enrollment forms, four sending birth forms, five sending exit forms, 15 sending encounter forms, and 10 sending group encounter forms. By May, 50 programs sent enrollment, 37 sent birth, 33 sent exit, 41 sent encounters, and 13 sent group encounter forms. Many of the programs that sent only minimal contact information and may not have sent any forms other than enrollment or encounter forms at first were CHW or Other programs, which may have seen participants only once or twice in public settings. Some of these programs did not collect data at all before this project, and they were able to slowly increase the number of questions they asked over time. Many programs still do not collect every type of form and skip questions, but there were vast improvements throughout the first year of the project.

#### Data analysis

Data created for the OEI reports and dashboard were analyzed using statistical software. Descriptive statistics that include counts, means, and confidence intervals, including bootstrapped estimations of these statistics, were calculated using STATA or Tableau. We also report inferential statistics for some of the metrics.

### Preliminary response counts and rates

The data model created by our research team has been leveraged to generate reports for key stakeholders and provide them with data visualizations using Tableau dashboards. Table 5 presents a summary of the proportion of data collection by mode for all of the OEI CBOs. Initial collection efforts suggest that CBOs are most comfortable reporting data through CCS or CBO Excel sheet. A combined 32% of CBOs reported through the Qualtrics system and validated Excel sheet modes. The paper form was the least used mode; 11% of CBOs used this mode to report their data.

**Table 5.** Counts and proportions of responses by data collection mode

| Data Collections Modes | Response Count (%) |
| --- | --- |
| CCS or CBO Excel format | 26 (41%) |
| OEI system (Qualtrics) | 12 (19%) |
| OCHIDS | 11 (17%) |
| OEI system (validated Excel file) | 8 (13%) |
| OEI system (paper form) | 7 (11%) |
| Total | 64 (100%) |

## Discussion

Data collection continues to be one of the biggest challenges for community-based organizations [28]. The OEI data collection system our research team developed is the foundational platform for a robust public health surveillance system to support interventions focused on infant mortality in Ohio. Even when the CBOs have systems in place, however, there are data points that are difficult to collect for less intensive program types. In our case, programs that meet occasionally in public settings or exist mainly to refer people to other services often see participants only once or twice. They also do not have time to collect extensive data during the encounters, nor has a trusting relationship been built with participants. There is more complete data collection for CenteringPregnancy and Home Visiting programs, so they may collect more complete demographic and risk factor data. Notwithstanding, CBOs that attempt to collect all or almost all of the variables in the OEI data collection system have missing data. Variables frequently missing information in the OEI data repository include data about the other biological parent, certain risk factors, and prenatal care attendance. This missing information is not systematic in nature, and such data quality concerns have motivated continuous changes to the data collection system. One approach has been to reword questions to provide clarity to items that are complex or difficult to understand.

Successes experienced in the first year of data collection included the ability to match to state records and improvements in data quality. Almost a third of participants were matched to birth records at the end of the first year. The lag in birth record reporting, the enrollment of participants early in pregnancy throughout the data collection period, and the collection of data about some participants who were not pregnant at enrollment or experienced pregnancy loss were factors that affected the ability to have all the records matched. Over time, the percentage linked to birth records will improve. In addition, over 60% were matched to Medicaid records. As there is not a requirement that participants receive Medicaid to participate in these programs, some participants may be uninsured, on private insurance, or on insurance associated with school or their parents. Finally, as more CBOs collected data and received support through PDSA cycles, the quality of data collection improved. Many CBOs who were not collecting sufficient data before this project worked with the research team to increase the number of variables collected. Allowing CBOs to submit data in multiple formats, provisioning reporting materials, and training on reporting data helped increase the number of programs submitting data and the completeness of this data. Despite some reluctance to add many new variables to data collection at needs assessment, the programs were able to comply with data collection because of the support they received and their desire for evaluation.

The OEI data collection system has been developed to assess statewide infant mortality prevention efforts, as there are many programs attempting to effect change but little coordination among programs. Although there are national reporting requirements for certain infant mortality prevention programs, including Nurse Family Partnership and CenteringPregnancy, there is little comparison of programs or compiling of program data. By collecting data for multiple kinds of programs in Ohio, the effects of any infant mortality prevention efforts can be assessed, and programs of similar types can be compared. Infant mortality is a rare outcome that is difficult to assess statistically, leading to difficulties assessing program success for small individual programs [9]. Compiling and harmonizing data across multiple small programs will allow for comparisons of birth outcomes both throughout the state and among program types to be more feasible, particularly for quantitative assessments where statistical power is needed. For example, if there is an infant mortality rate of 5 per 1,000 in a 400 person CenteringPregnancy programmatic intervention, it may not be a statistically significant reduction compared to a general population with a rate of 6 per 1,000; however, if the population is 20,000 across multiple CenteringPregnancy programs, the differences in infant mortality rates may reach statistical significance because of an increase in statistical power. The results of these comparisons will help highlight which program types and components have the most potential for improving birth outcomes, allowing funding and program decisions to be made within Ohio. These evaluations may also help other states adopt similar data collection systems and compare efforts among states.

Data collection for community programs also has the potential to lead to coordinated efforts with health care providers. Infant mortality has multifaceted risks that cannot be addressed through the medical model alone. Many of the environmental, social, and behavioral risks that participants may experience are addressed by community programs through provision of referrals to other services, provision of child care supplies, social support, and education that is not standard for prenatal care in a medical setting [29-32]. Participating in these programs, if shown to be successful at improving maternal and child health, could be encouraged by medical providers. Coordination with community programs could improve health care quality even more if there was potential for information exchange among programs and providers, as participants may report and focus on different risks with a community health worker than they do with their doctors. Additionally, systematic collection of risk factors incorporates these critical social determinants of health to present a comprehensive picture about a patient to the provider and enhances the overall value of care delivered to the patient [33-34]. This type of information would include aggregate information about patients’ environments and the risk factors they experience.

Improvements in public health require information systems for surveillance and effective program implementation and impact, along with partnerships that build public support [27]. Because of the various risks for infant mortality across preconception and throughout pregnancy, infant mortality prevention efforts need to collaborate to achieve lasting change at a large scale, not just in Ohio [3, 4]. Statewide efforts of this nature to standardize data collection could be used as a template for enhanced nationwide infant mortality efforts. The template, moreover, can also be extended to other community services, such as those that target children with disabilities or are related to people with mental and behavioral health conditions.

## Conclusion

There is a need to develop an ecosystem of tools to effectively gather data from program participants, community members, community-based organizations, and local and state authorities to facilitate a greater understanding of how public health interventions impact the communities they serve. There also is a need for participant-facing tools to collect information about service quality and patient-reported outcomes without having to report through a community health worker or other intermediaries. Potential data collection tools include smartphone applications, text messaging-based data collection, and interactive voice activated applications. In addition, there is a need for organization-facing tools that report information back to organizations about their performance relative to other organizations and information about whether there are gaps in service. Currently, the Tableau dashboards are used by the program sponsors, and some of these results may be reported back to CBOs; however, these organizations do not have control over what data they see. Systems may be needed to provide regular feedback and data access for analysis for individual CBOs. Organizational engagement will be imperative moving forward to develop more efficient systems of bi-directional reporting.

The OEI data collection infrastructure our research team designed and deployed is still in its early stages of system maturity. Our research team continues to learn from its implementation and use as the system evolves over time. The system, although requiring more effort from CBOs, is already demonstrating signs of collective action at the local and state levels to better coordinate and share information on how to best utilize programmatic resources to reduce infant mortality and its associated disparities in across Ohio.

## Data Availability

Data used in this study was reviewed and approved by the Ohio Department of Medicaid and is not publicly available.

## Acknowledgements

The authors would like to thank all the staff members of the Ohio Colleges of Medicine Government Resource Center and the Ohio Department of Medicaid for their input during the development of our infrastructure. We are also grateful for their feedback on our manuscript. We also acknowledge the financial assistance from the Ohio Department of Medicaid under Task Order ODM202012, which supported the building of our infrastructure.

## Conflicts of Interest

None declared.

## Abbreviations

CBO: Community-Based Organization
CCS: Care Coordination Systems
OCHIDS: Ohio Comprehensive Home Visiting Integrated Data System
ODM: Ohio Department of Medicaid
OEI: Ohio Equity Institute
PDSA: Plan-Do-Study-Act

## Additional Files

Additional File 1.docx

Variable list

This file includes the complete list of variables measured using the enrollment, birth, exit and encounter forms used as part of this study.

Additional File 2.docx

OEI Data Dictionary

This file includes the complete data dictionaries for the enrollment, birth, exit, and encounter forms used as part of this study.

## Figures

Readers may contact the corresponding author to request access to this image.

## Notes

### Competing Interest Statement

The authors have declared no competing interest.

### Author Declarations

This study was deemed exempt for review by The Ohio State University IRB.

