## Supplementary Table for "Co-establishing an infrastructure for routine data collection to address disparities in infant mortality: Planning and implementation"

Supplementary Table 1. These variables were measured using the enrollment form.

|  | Variable | Description |
| --- | --- | --- |
| **Demographics and Identification** |  |  |
|  | First and Last Name | Participant and other biological parent |
|  | Social Security Number or Medicaid ID | Participant |
|  | Date of Birth | Participant and other biological parent; age calculated from this variable |
|  | Race/Ethnicity | Participant and other biological parent; census standard is used unless there is an alternative preference |
|  | Marital Status | Participant marital status |
|  | Household Size | Count of the number of people (adults plus children) who live in the house with the participant |
| **Behavioral Risk Factors** |  |  |
|  | Alcohol Usage | Participant or anyone in their home drinking alcohol regularly and frequency of use |
|  | Maternal Diet/Folic Acid Consumption | Participant intake of prenatal vitamins |
|  | Smoking/Smoking Cessation | Participant or anyone in their home smoking regularly and how often |
|  | Substance Abuse | Participant using any substances regularly and frequency of use; whether friends or family use substances |
|  | Depression | Participant history of depression diagnosis or treatment |
|  | Stress | Participant experience with significant stress in past month |
|  | Social Support | Participant access to emotional and practical support from family and peers |
| **Existing Prenatal Care** |  |  |
|  | Self-reported Weeks Gestation at Enrollment | Count of weeks participant is pregnant upon enrollment to program |
|  | Self-reported Treatment with Progesterone | Participant receipt of progesterone treatment during current pregnancy |
| **Social Programmatic Assessment** |  |  |
|  | Referral Source | Referral source of participant for program enrollment |
|  | Enrollment in Food Stamps | Participant receipt of food from the Supplemental Nutrition Assistance Program (SNAP) |
|  | Welfare Use | Participant receipt of money for disability, unemployment, or Temporary Assistance for Needy Families (TANF) |
|  | Program Use | Participant enrollment in any other social or community programs |
| **Environmental Context** |  |  |
|  | Economic Self-Sufficiency | Participant need for monetary aid and ability to pay bills without help |
|  | Nutritional Sufficiency | Participant access to sufficient food and nutrition |
|  | Housing/Housing Instability | Participant access to consistent place to live and type of housing |
|  | Home Environment/Safety | Participant ability to live in a safe place (home has heat, water, electricity, and no pests/irritants/hazards) |
|  | Employment Status | Participant employment, academic, disability, or other employment status |
|  | Transportation Access | Participant access to a vehicle, bus, or other transportation and ability to get to between places |

Supplementary Table 2. These variables were measured using the encounter form.

|  | Variable | Description |
| --- | --- | --- |
| **Demographics and Identification** |  |  |
|  | First and Last Name | Participant |
|  | Date of Birth | Participant |
| **Program Evaluation** |  |  |
|  | Referrals Given to Services | Social or medical services a participant was referred to at a visit |

Supplementary Table 3. These variables were measured using the birth form

|  | Variable | Description |
| --- | --- | --- |
| **Demographics and Identification** |  |  |
|  | First and Last Name | Participant and infant |
|  | Social Security Number or Medicaid ID | Participant and infant |
|  | Infant Date of Birth | Infant |
|  | Infant Race/Ethnicity | Infant |
| **Behavioral Risk Factors** |  |  |
|  | Breastfeeding Rate/Feeding Practices | How infant is being fed |
|  | Infant Sleep Location/Position | Type of surface infant sleeps on and sleep position/environment |
| **Existing Prenatal Care** |  |  |
|  | Self-reported Treatment with Progesterone | Participant receipt of progesterone treatment |

Supplementary Table 4. These variables were measured using the exit form.

|  | Variable | Description |
| --- | --- | --- |
| **Demographics and Identification** |  |  |
|  | First and Last Name | Participant and infant |
|  | Social Security Number or Medicaid ID | Participant and infant |
| **Existing Care** |  |  |
|  | Immunization | Adherence to recommended immunization schedule for infants in the first year of life (for CBOs that have follow-up through one year) |
|  | Attendance at Postpartum Visits | Count of postpartum visits a participant attends |
|  | Well-Child Visits | Count of well-child visits attended by a participant and child |
|  | Emergency Room/Urgent Care Visits | Count of any visits to the emergency room or urgent care for a participant or child |
| **Social Programmatic Assessment** |  |  |
|  | Program Use | Participant enrollment in any other community programs |
|  | Enrollment in Food Stamps | Participant receipt of food from the Supplemental Nutrition Assistance Program (SNAP) |
|  | Welfare Use | Participant receipt of money for disability, unemployment, or Temporary Assistance for Needy Families (TANF) |
|  | Employment Status | Participant employment, academic, disability, or other employment status |
| **Environmental Context** |  |  |
|  | Child Care | The child care arrangements if a parent cannot be home |
|  | Housing/Housing Instability | Participant access to consistent place to live and type of housing |
|  | Father Engagement | Status of father’s involvement in parenting/caring for the child |
|  | Home Environment/Safety | Participant and infant ability to live in a safe place (home has heat, water, electricity, and no pests/irritants/hazards) |
|  | Nutritional Sufficiency | Participant and infant access to sufficient food and nutrition |
