## Supplementary Table for "Co-establishing an infrastructure for routine data collection to address disparities in infant mortality: Planning and implementation"

Supplementary Table 5. OEI Data Dictionary: Enrollment Form

| Full question text | Answer choices |
| --- | --- |
| On what date did the participant enroll in the program? | mm/dd/yyyy format |
| What is the participant's birth date? | mm/dd/yyyy format |
| Participant's identification information: First name | Any text, maximum 14 characters |
| Participant's identification information: Last name | Any text, maximum 18 characters |
| Participant's identification information: Social Security number | Must be 9 digits |
| Participant's identification information: Medicaid ID number | Must be 12 digits |
| Participant's identification information: Gender | Female; Male |
| Participant's identification information: Street address | Any text, maximum 35 characters |
| Participant's identification information: City | Any text, maximum 13 characters |
| Participant's identification information: ZIP code | Must be 5 digits |
| Participant's identification information: Phone number | Must be 10 digits |
| How did the participant learn about this program? | Friend or family member; Medical provider; Other prenatal or infant care program; Social/governmental program; Advertisements in the community; Other (please specify) |
| How did the participant learn about this program? - Other (please specify) - Text | Textbox - can enter any information on how participant learned about program, maximum 15 characters |
| Participant's employment status (please check all that apply): | Employed full time; Employed part time; Unemployed, receiving assistance; Unemployed, not receiving assistance; Enrolled in school; Disabled |
| Participant's marital status: | Married; Widowed; Divorced; Separated; Single/never married; Not married but living with partner |
| Current relationship between participant and other biological parent: | Married; Widowed; Divorced; Separated; Never married |
| Living status between participant and other biological parent: | Living together; Not living together |
| Other biological parent's birth date: | mm/dd/yyyy format |
| Participant's race/ethnicity (please check all that apply): | White; Black or African American; Hispanic or Latino; Asian; Native Hawaiian or Pacific Islander; American Indian or Alaska Native; Other (please specify) |
| Participant's race/ethnicity (please check all that apply): - Other (please specify) - Text | Textbox - can enter any information on race, maximum 15 characters |
| Other biological parent's race/ethnicity (please check all that apply) | White; Black or African American; Hispanic or Latino; Asian; Native Hawaiian or Pacific Islander; American Indian or Alaska Native; Other (please specify) |
| Other biological parent's race/ethnicity (please check all that apply) - Other (please specify) - Text | Textbox - can enter any information on race, maximum 15 characters |
| Other biological parent's identification information: - First name | Any text, maximum 14 characters |
| Other biological parent's identification information: - Last name | Any text, maximum 18 characters |
| Other biological parent's identification information: - Street address | Any text, maximum 35 characters |
| Other biological parent's identification information: - City | Any text, maximum 13 characters |
| Other biological parent's identification information: - ZIP code | Must be 5 digits |
| Other biological parent's identification information: - Phone number | Must be 10 digits |
| How many total adults live in the same household as the participant (including the participant)? | Allows 2 digits |
| How many total children live in the same household as the participant? | Allows 2 digits |
| What kind of housing does the participant have? | Live in house/apartment owned by participant; Live in house/apartment owned by family/friends; Live in rented house/apartment; Live in shelter/group home; Public housing; Homeless; Other (please specify) |
| What kind of housing does the participant have? - Other (please specify) - Text | Textbox - can enter other kinds of housing, maximum 15 characters |
| Please check any home safety issues the participant is experiencing. | No working smoke detectors; Firearms or weapons in home; Smell of gas/mildew/mold; Pests suspected/present; Smoking in house; Windows/doors do not lock appropriately; Garbage/clutter/unclean environment; Drugs/chemicals/cleaning supplies within reach |
| Participant’s primary method of transportation: | Own car; Bus; Taxi; Walk; Friend’s/family car; Other (please specify) |
| Participant’s primary method of transportation: - Other (please specify) – Text | Textbox – can enter any information on transportation, maximum 15 characters |
| Is the participant currently enrolled in any of the following public assistance programs? (Please check all that apply.) | Women, Infants and Children (WIC); Supplemental Nutrition Assistance Program (SNAP/food stamps); Temporary Assistance for Needy Families (TANF); Disability or unemployment; Other prenatal/infant health program; Housing assistance program; Child care program; Food assistance program; Exercise/health promotion program; Education/employment assistance program; Mental health/substance abuse program; Don’t know; Other (please specify) |
| Is the participant currently enrolled in any of the following public assistance programs? (Please check all that apply.) – Other (please specify) – Text | Textbox – can enter any information on programs, maximum 15 characters |
| Is the participant financially stable (able to pay their bills without any monetary aid or help)? | Yes; No |
| Does the participant have access to adequate food? | Yes; No |
| Does the participant have current depression or a history of depression diagnosis or treatment? | Yes; No |
| In the past month, did the participant feel they could not control important things in their life? | Yes; No |
| Is there at least one person the participant can discuss their thoughts and feelings with? | Yes; No |
| Does the participant or anyone in their household smoke? (If no, skip to #27.) | The participant smokes but nobody else in the household smokes; At least one member of the household smokes but the participant does not; Both the participant and at least one other person in the household smoke; No one in the household smokes |
| If the participant smokes, number of cigarettes smoked per day: | Less than one per day; 1-5; 6-10; 11-15; 16-20; More than 20 per day |
| If others in the household smoke, number of cigarettes smoked per day: | Less than one per day; 1-5; 6-10; 11-15; 16-20; More than 20 per day |
| In the past month, has the participant drank any alcohol? | Yes; No |
| In the past six months, has the participant used any illegal substances? (If no, skip to #30.) | Yes; No |
| If yes to #28, how frequently does the participant use controlled substances? | More than once per day; Once per day; A few times per week or less; A few times per month or less; Only on occasion/less than once per month |
| Do any of the participant’s friends or family members have problems with alcohol or other drug use? | Yes; No |
| Participant's weeks of gestation at enrollment in program: | Allows 2 digits |
| Participant's weeks of gestation at enrollment in program: Checkbox | Check to indicate infant has already been born |
| How many prenatal care visits has the participant had prior to enrollment? | Allows 2 digits |
| Did the participant have any prenatal visits in her first trimester (weeks 1-12)? | Yes; No |
| Has the participant received treatment with progesterone during this pregnancy? | Yes; No |
| Does the participant take folic acid/vitamins? | Yes; No |
| Is transportation a barrier to the participant attending prenatal care appointments? | Yes; No |

Supplementary Table 6. OEI Data Dictionary: Birth Form

| Full question text | Answer choices |
| --- | --- |
| On what date is this form being filled out? | Mm/dd/yyyy format |
| What is the participant’s birth date? | Mm/dd/yyyy format |
| Participant’s identification information: First name | Any text, maximum 14 characters |
| Participant’s identification information: Last name | Any text, maximum 18 characters |
| Participant’s identification information: Social Security number | Must be 9 digits |
| Participant’s identification information: Medicaid ID number | Must be 12 digits |
| Participant’s identification information: Gender | Female; Male |
| Participant’s identification information: Street address | Any text, maximum 35 characters |
| Participant’s identification information: City | Any text, maximum 13 characters |
| Participant’s identification information: ZIP code | Must be 5 digits |
| Participant’s identification information: Phone number | Must be 10 digits |
| Infant’s identification information: First name | Any text, maximum 14 characters |
| Infant’s identification information: Last name | Any text, maximum 18 characters |
| Infant’s identification information: Social Security number (if known) | Must be 9 digits |
| Infant’s identification information: Medicaid ID number (if known) | Must be 12 digits |
| Infant’s identification information: Sex | Female; Male |
| Is the infant’s address the same as the participant’s? | Yes; No |
| Infant’s identification information: Street address | Any text, maximum 28 characters |
| Infant’s identification information: City | Any text, maximum 13 characters |
| Infant’s identification information: ZIP code | Must be 5 digits |
| On what date was the infant born? | Mm/dd/yyyy format |
| Was this a multiple birth? | No; Yes, twins; Yes, triplets |
| What is the infant’s race/ethnicity? (Please check all that apply.) | White; Black or African American; Hispanic or Latino; Asian; Native Hawaiian or Pacific Islander; American Indian or Alaska Native; Other |
| What is the infant’s race/ethnicity? (Please check all that apply.) – Other (please specify) – Text | Textbox – can enter any information on race, maximum 15 characters |
| How is the infant being fed? | Breastfeeding only; Some breastfeeding, some formula; Formula only |
| Where does the infant sleep? (Please check all that apply.) | Adult bed; Crib; Sofa/couch/chair; Car seat; Floor; Bassinet; Pack ‘n Play; Other (please specify) |
| Where does the infant sleep? (Please check all that apply.) – Other – Text | Textbox – can enter any other sleep surface, maximum 15 characters |
| What position is the infant put to sleep in most frequently? | On their back; On their side; On their stomach |
| Does the infant ever share a sleeping surface with any other people or a pet? | Yes; No |
| Did the participant receive treatment with progesterone during this pregnancy? | Yes; No |
| How many total prenatal visits did the participant attend during this pregnancy? | Allows 2 digits |

Supplementary Table 7. OEI Data Dictionary: Exit Form

| Full question text | Answer choices |
| --- | --- |
| On what date did the participant exit the program? | mm/dd/yyyy format |
| What is the participant's birth date? | mm/dd/yyyy format |
| Participant's identification information: First name | Any text, maximum 14 characters |
| Participant's identification information: Last name | Any text, maximum 18 characters |
| Participant's identification information: Social Security number | Must be 9 digits |
| Participant's identification information: Medicaid ID number | Must be 12 digits |
| Participant's identification information: Gender | Female; Male |
| Participant's identification information: Street address | Any text, maximum 35 characters |
| Participant's identification information: City | Any text, maximum 13 characters |
| Participant's identification information: ZIP code | Must be 5 digits |
| Participant's identification information: Phone number | Must be 10 digits |
| Infant's identification information: First name | Any text, maximum 14 characters |
| Infant's identification information: Last name | Any text, maximum 18 characters |
| Infant's identification information: Social Security number (if known) | Must be 9 digits |
| Infant's identification information: Medicaid ID number (if known) | Must be 12 digits |
| Infant's identification information: Sex | Female; Male |
| Is the infant's address the same as the participant's? | Yes; No |
| Infant's identification information: Street address | Any text, maximum 28 characters |
| Infant's identification information: City | Any text, maximum 13 characters |
| Infant's identification information: ZIP code | Must be 5 digits |
| On what date was the infant born? | mm/dd/yyyy format |
| Was this a multiple birth? | No; Yes, twins; Yes, triplets |
| What kind of housing does the participant have? | Live in house/apartment owned by participant; Live in house/apartment owned by family/friends; Live in rented house/apartment; Live in shelter/group home; Public housing; Homeless; Other (please specify) |
| What kind of housing does the participant have? - Other (please specify) - Text | Textbox - can enter other kinds of housing, maximum 15 characters |
| Please check any home safety issues the participant is experiencing. | No working smoke detectors; Firearms or weapons in home; Smell of gas/mildew/mold; Pests suspected/present; Smoking in house; Windows/doors do not lock appropriately; Garbage/clutter/unclean environment; Drugs/chemicals/cleaning supplies within reach |
| Participant's Employment Status (please check all that apply): | Employed full time; Employed part time; Unemployed, receiving assistance; Unemployed, not receiving assistance; Enrolled in school; Disabled |
| Is the participant currently enrolled in any of the following public assistance programs? (Please check all that apply.) | Women, Infants and Children (WIC); Supplemental Nutrition Assistance Program (SNAP/food stamps); Temporary Assistance for Needy Families (TANF); Disability or unemployment; Other prenatal/infant health program; Housing assistance program; Child care program; Food assistance program; Exercise/health promotion program; Education/employment assistance program; Mental health/substance abuse program; Don't know; Other (please specify) |
| Is the participant currently enrolled in any of the following public assistance programs? (Please check all that apply.) - Other (please specify) - Text | Textbox - can enter any information on programs, maximum 15 characters |
| Does the participant have access to adequate food? | Yes; No |
| How many well-child medical visits did the participant and child attend to check on the child's health? | Allows 2 digits |
| How many ER/Urgent care visits were attended for the child or participant since the child's birth? | Allows 2 digits |
| If the answer to #13 was more than zero, what were the reasons for the visit/visits? | Textbox - can describe reasons for ER/urgent care, no character restriction |
| Has the child received recommended immunizations? | Yes; No |
| When the participant is away from home, where does the child or children go for child care? | Family; Friends; Licensed child care provider; Other (please specify) |
| When the participant is away from home, where does the child or children go for child care? - Other (please specify) - Text | Textbox - can enter place of child care, maximum 15 characters |
| How many postpartum medical visits did the mother attend to check on her health? | Allows 2 digits |
| Is the father involved in the care of the infant? | Yes; No |

Supplementary Table 8. OEI Data Dictionary: Encounter Form

| Full question text | Answer choices |
| --- | --- |
| On what date did this encounter occur? | Mm/dd/yyyy format |
| What is the participant’s birth date? | Mm/dd/yyyy format |
| Participant’s identification information: First name | Any text, maximum 14 characters |
| Participant’s identification information: Last name | Any text, maximum 18 characters |
| Participant’s identification information: Street address | Any text, maximum 28 characters |
| Participant’s identification information: Gender | Female; Male |
| Participant’s identification information: City | Any text, maximum 13 characters |
| Participant’s identification information: ZIP code | Must be 5 digits |
| Participant’s identification information: Phone number | Must be 10 digits |
| What is the next planned contact date? | Mm/dd/yyyy format |
| Was the participant referred to any additional programs or services? | Yes; No |
| If yes to #5, please list any additional programs or services the participant was referred to. | Textbox – list what participant was referred to here |
| *Encounter form – linkage information* |  |
| Form administrator’s first name | Any text, maximum 14 characters |
| Form administrator’s last name | Any text, maximum 18 characters |
| Organization/program name | Any text, maximum 20 characters |
| Phone number for questions | Must be 10 digits |
